# Downregulation of cancer hallmarks and immune check-points in patients with glioblastoma following a short course of the pro-oxidant combination of Resveratrol and Copper

**DOI:** 10.1101/2025.02.19.25322384

**Authors:** Chaitra Bandiwadekar, Naorem Leimarembi Devi, Aliasgar V Moiyadi, Vikas Singh, Prakash Shetty, Sridhar Epari, Harshali Tandel, Roohi Yelukar, Disha Poojary, Gorantla V Raghuram, Snehal Shabrish, Pratik Chandrani, Indraneel Mittra

## Abstract

**Background:** We investigated a novel therapeutic approach to glioblastoma (GBM) that targets cell-free chromatin particles (cfChPs) that are released from dying GBM cells and aggravate the oncogenic phenotype of living GBM cells. cfChPs can be deactivated by oxygen radicals (OR) generated upon admixing the nutraceuticals Resveratrol (R) and Copper (Cu). Oral administration of R-Cu leads to generation of OR which are readily absorbed to deactivate cfChPs.

**Patients and Methods:** Ten patients with glioblastoma awaiting surgery were administered tablets containing 5.6mg of Resveratrol (R) and 560ng of Copper (Cu) four times a day for an average of 11.6±5.37 days. Another ten patients who did not receive R-Cu acted as controls. A biopsy of the tumour tissues was taken at operation for analysis using confocal microscopy, immunofluorescence and transcriptome sequencing.

**Results:** Confocal microscopy of tumour sections revealed copious presence of cfChPs in the tumour microenvironment (TME) that had been released from dying GBM cells. R-Cu treatment led to deactivation / eradication of cfChPs. Eradication of cfChPs from TME led to a highly significantly reduction in Ki-67 and nine hallmarks of cancer, six immune check-points and three stem cell biomarkers. Transcriptome sequencing detected marked upregulation of pro-apoptotic and down-regulation of anti-apoptotic genes. Also detected was down-regulation of PVRIG-2P, a homologue of immune checkpoint receptor PVRIG which is a functional analogue of PD-L1.

**Conclusion:** The results of our study suggest that oral administration of a non-toxic combination of small quantities of the commonly used nutraceuticals R and Cu can have a profound effect in attenuating the aggressive phenotype of GBM. They also suggest that cfChPs are the key activators of cancer hallmarks, immune checkpoints and cancer stemness in GBM. Further studies are required to investigate whether prolonged treatment with R-Cu may induce the tumour to adopt a benign phenotype.

**Trial registration:** ClinicalTrials.gov identifier: CTRI/2020/10/028476 (https://ctri.nic.in/Clinicaltrials/pmaindet2.php?EncHid=NDY2Mzc=&Enc=&userName=Al iasgar)

**Graphical Abstract:** 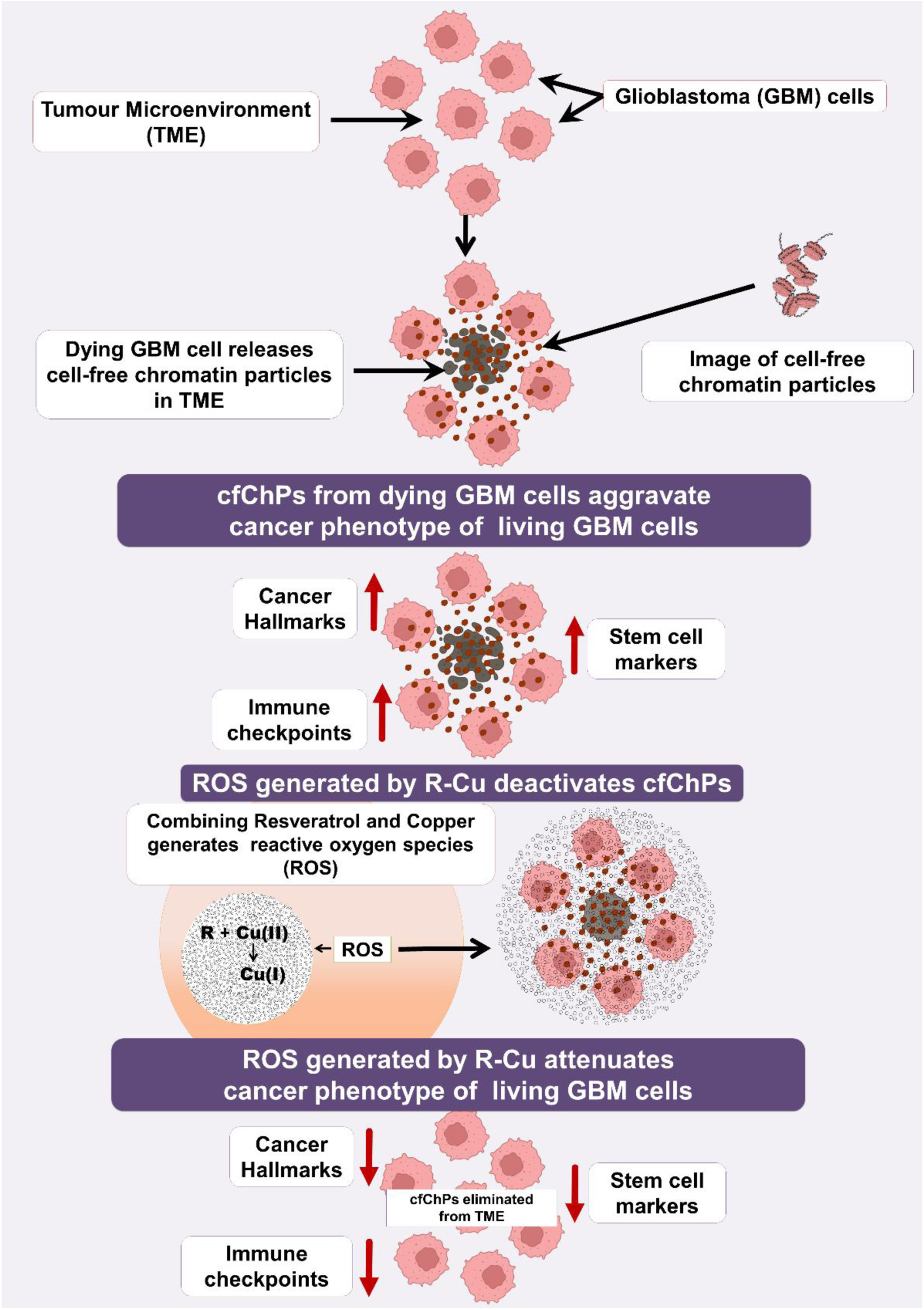

Attenuation of malignant phenotype of glioblastoma by oxygen radicals (ROS) generated by combining Resveratrol and Copper.

## Introduction

Glioblastoma (GBM) is a highly aggressive tumour with poor prognosis. Despite intensive combination therapies involving surgery, radiotherapy and chemotherapy, glioblastoma (GBM) remains an incurable disease with a median survival of 15 months (Thakkar et al., 2014). Novel therapeutic approaches that are less traumatic and non-toxic are needed. We previously reported that cell-free chromatin particles (cfChPs) that are released from dying cancer cells can be readily internalized by bystander living cells leading to the activation of two crucial hallmarks of cancer viz. dsDNA breaks (genome instability) and inflammation (Mittra et al., 2017b). Given that the concurrent induction of DNA damage and inflammation is a powerful trigger for oncogenic transformation (Balkwill et al., 2005; Hanahan and Weinberg, 2011), these findings suggested that cfChPs released into the tumour microenvironment (TME) from dying cancer cells may aggravate the malignant phenotype of the surviving cancer cells. We have also reported that the activation of DNA damage and inflammation can be abrogated by concurrent treatment with a combination of the nutraceuticals resveratrol (R) and copper (Cu) which can deactivate the cfChPs via the medium of oxygen radicals (Kirolikar et al., 2018; Mittra et al., 2017b). It was first reported by Fukuhara et. al. that R functions as a catalyst to reduce Cu (II) to Cu (I) resulting in the generation of oxygen radicals which can cleave plasmid pBR322 DNA (Fukuhara et al., 2006; Fukuhara and Miyata, 1998). We have extended these findings by demonstrating that oxygen radicals produced after admixing R and Cu (R-Cu) can deactivate extracellular cfChPs by breaking down their DNA component (Mittra, 2024; Subramaniam et al., 2016). When R-Cu is taken orally, oxygen radicals are generated in the stomach, which are readily absorbed and have systemic effects in the form of deactivation of extracellular cfChPs. We have reported, in pre-clinical and clinical studies, that R-Cu can reduce toxicities associated with chemotherapy (Agarwal et al., 2022; Mittra et al., 2017a; Ostwal et al., 2022) and radiotherapy (Kirolikar et al., 2018), can minimize fatality from sepsis (Mittra et al., 2020b, 2020a) and downregulate multiple biomarkers of ageing and neurodegeneration (Pal et al., 2022). No toxic side effects attributable to R-Cu were reported in any of the above pre-clinical or clinical studies.

In the present study we tested the hypothesis that oral administration of R-Cu would lead to deactivation of cfChPs that are released into the TME from dying GBM cells and prevent them from aggravating the malignant phenotype of the surviving cancer cells. Our earlier clinical studies had indicated the optimum dose of R and Cu to be 5.6 mg and 560 ng respectively (Agarwal et al., 2022; Mittra et al., 2020a; Ostwal et al., 2022; Pilankar et al., 2022). These doses of R and Cu were combined in a bi-layered tablet formulation and used in this study. The tablets were provided for research purposes only by Inventia Healthcare Limited, Mumbai (Batch no. SC2008, Expiry date: October 2024).

## Results

### R-Cu treatment eradicates cfChPs from TME

Confocal microscopy following fluorescent immune staining of FFPE sections using anti-DNA and anti-histone H4 revealed copious presence of cfChPs in TME of the untreated samples tissues (Fig. 1A, upper panel). Under normal circumstances the DNA (red) and histone H4 (green) fluorescent signals would be expected to be restricted to the nuclei (Fig. 1A). However, we detected extensive presence of red and green fluorescent signals outside the nuclei which upon superimposing the red and green images appeared as yellow fluorescent signals representing cfChPs. This indicated that the extensive apoptotic cell death had resulted in copious release of cfChPs in TME. The yellow fluorescent particles were virtually absent in the R-Cu treated samples indicating that R-Cu treatment had deactivated / eliminated the cfChPs from TME (Fig. 1A, lower panel). This finding is quantitatively depicted in the form of histograms which indicated a reduction in extra-nuclear mean fluorescent intensity (MFI) in the R-Cu treated samples compared to the controls (p < 0.01).

**Figure 1:**
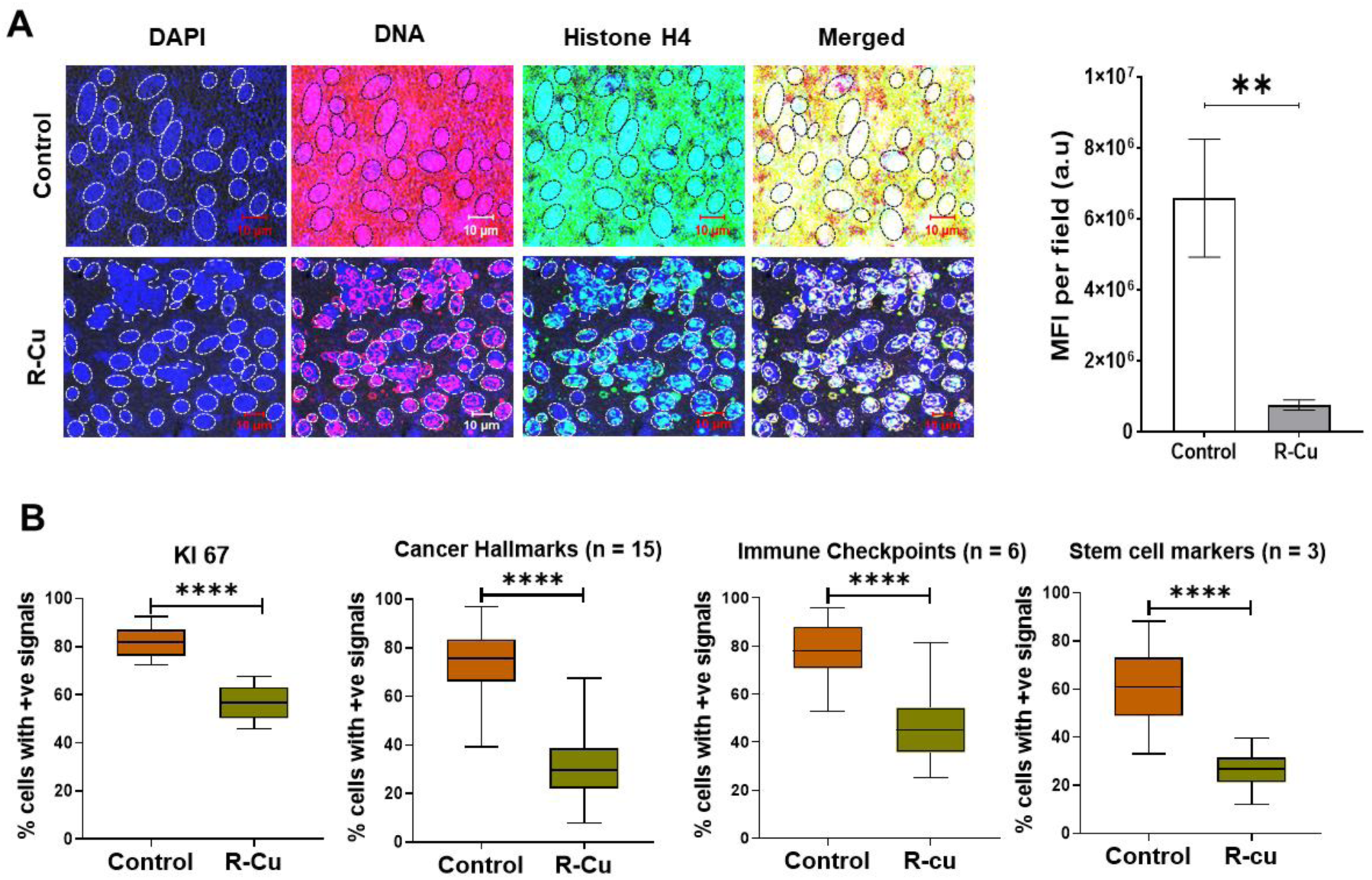
Representative confocal microscopy images of tumour sections to detect cfChPs in TME, and Immunofluorescence results of Ki-67, and combined results of 15 hallmarks of cancer, 6 immune check-points and 3 stem cell markers in control and R-Cu treated GBM patients. **A.** Representative fluorescence immuno-staining and confocal microscopy images of tumour sections of GBM samples stained with fluorescent antibodies against DNA and histone H4 and examined by confocal microscopy. Co-localizing DNA (red) and histone H4 (green) fluorescent signals generate yellow / white coloured particles that represent cfChPs. Numerous yellow / white fluorescent particles are seen outside the nucleus in the intra- or extracellular spaces in the control samples which are virtually eliminated following R-Cu treatment. Graphical representation of MFI of extra-nuclear cfChPs (right hand image). The nuclei were gated and MFI of yellow fluorescent signals in five randomly chosen confocal fields (∼50 cells per field) was estimated. The number of patient samples in control and R-Cu treated groups = 10 in each group. **B.** Boxplots representing Mean ± SEM of immunofluorescence results of Ki-67 and combined results of 15 hallmarks of cancer, 6 immune check-points and 3 stem cell markers. n = 10 patients each in control and R-Cu treated patients. ** = p < 0.01; **** = p < 0.0001.

### R-Cu treatment down-regulates cancer biomarkers

We analyzed by IF on four groups of biomarkers which included: 1) Ki-67; 2) fifteen biomarkers representing nine hallmarks of cancer; 3) six immune check-points and 4) three stem cell markers. Ki-67 labeling is routinely used in clinical practice as a measure of proliferative index of GBM (Xu et al., 2021). We observed highly significant reduction in Ki-67 levels in the R-Cu treated samples compared with untreated samples suggesting a downward shift in the histological grade of malignancy of the tumours (Mean ± SEM = 82.09 ± 2.03 vs 56.72 ± 2.33; p < 0.0001) (though the morphological features like quantum of necrosis/foci of microvascular proliferation did not show any significant change) (Fig. 1B, first panel). Representative IF images of Ki-67 in control and R-Cu samples is given in Fig. S1. The 15 biomarkers representing 9 hallmarks of cancer defined by Hanahan and Weinberg (Hanahan and Weinberg, 2011) that we analyzed are listed in Table S2. The combined results of percent cells with positive signals of the 15 biomarkers are represented as box-plot in Fig. 1B (second panel). We observed a highly significant reduction in the combined results of hallmarks of cancer in the R-Cu treated samples compared to untreated controls (p < 0.0001). Individual IF images and box-plot results of the 15 biomarkers in treated and untreated samples is given in Fig. S2. It can be observed from Fig. S2 that there was highly significant reduction in the levels of individual biomarkers in the R-Cu treated groups compared to controls in 13 / 15 cases (p < 0.0001). The only exception was ppP53 where the p value was 0.01. The immune check-points analyzed by IF were PD-1, PD-L1, TIM-3, NKG2A, CTLA-4 and LAG3. Here again the combined results demonstrated highly significant reduction of immune check-points in the R-Cu treated samples compared to untreated controls (p < 0.0001) (Fig. 1B, third panel). Individual IF images and box-plot results of the 6 immune check-points in treated and untreated samples is given in Fig. S3. In all cases except CTLA-4 the p value was < 0.0001. We confirmed by IF that the 5 immune check-points viz. PD-1, TIM-3, NKG2A, CTLA-4 and LAG3 co-localized with tumour infiltrating lymphocytes marked by CD3 indicating that these immune check-points were being expressed by tumour infiltrating lymphocytes (Fig. S4).

We next analyzed three stem cell markers viz. CD133, CD44 and SOX2. Of these, the first two are cancer specific while SOX2 is a general stem cell marker. The combined box-plot results revealed a highly significant reduction in the R-Cu treated samples compared to untreated controls (p < 0.0001) (Fig. 1B, fourth panel). Individual IF images and box-plot results of the 3 stem cell markers in treated and untreated samples is given in Fig. S5. In each case the p value was highly significant at p < 0.0001.

### RNA-seq analysis

#### Quality check

Out of the 20 samples, 6 samples from each group passed the quality check and were used for RNA-seq analysis. The 12 samples were sequenced for whole transcriptome analysis, with a median coverage of 42 million reads (ranging from 30-51 million) (Table S3). FastQC was used to evaluate mean per-base quality score and per-sequence quality score of RNA-seq data. We obtained median Phred quality score above 30, confirming the high quality of RNA-seq reads (Figs. S6A & S6B). Also, the boxplot of Cook’s distance distribution showed that all samples had a normalized distribution of gene expression (Fig. S6C).

#### Clear clustering of R-Cu transcripts

Whole transcriptome multidimensional scaling (MDS) analysis identified distinct separation between R-Cu from untreated samples (Fig. 2A). The plot indicated that samples formed a cluster (teal), showing closeness of genotype upon R-Cu treatment clearly separating them from control samples. The separation of R-Cu treated samples from control sample indicated activation of a novel transcriptional circuitry upon R-Cu treatment.

**Figure 2:**
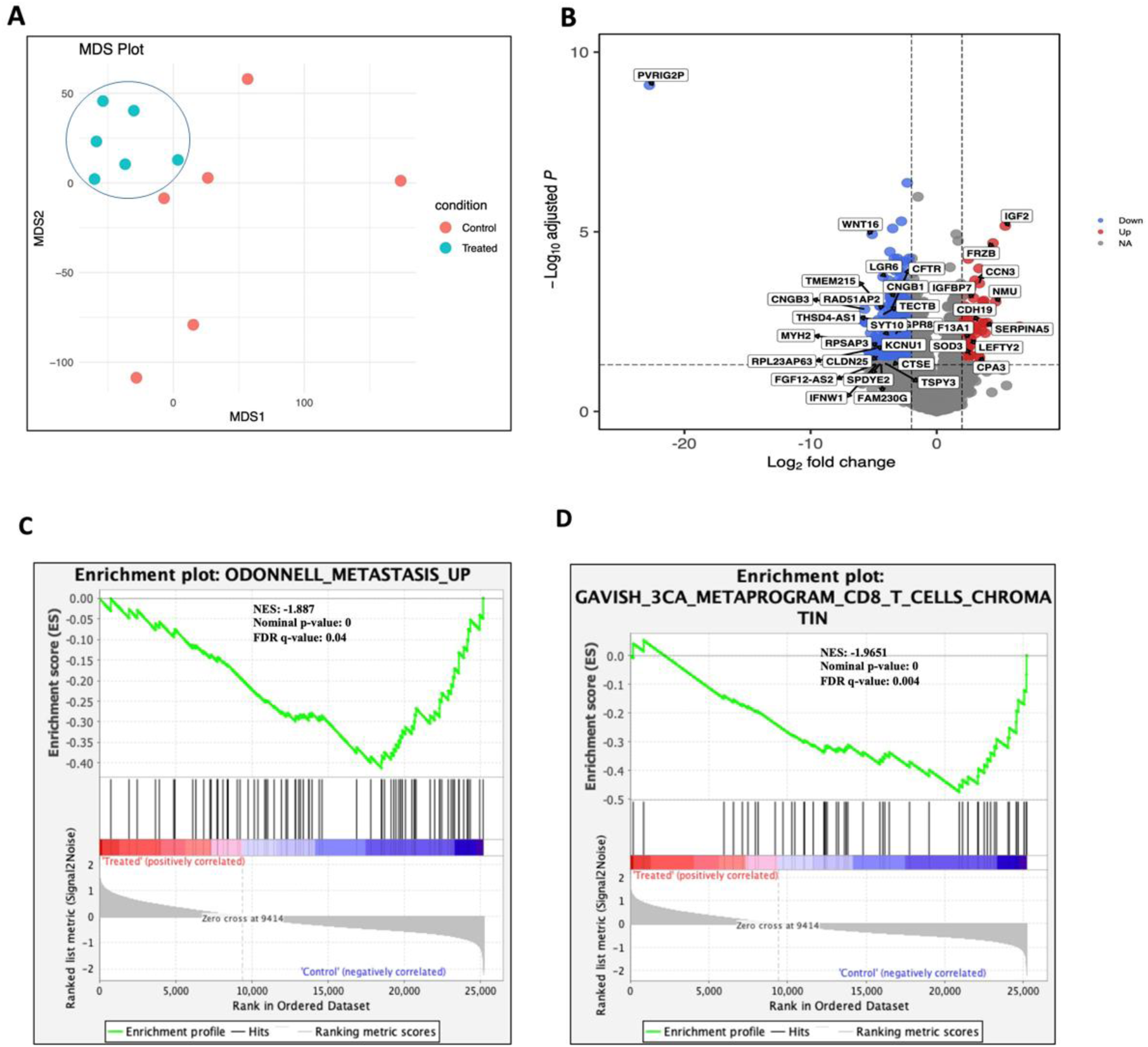
RNA-seq analysis of glioblastoma patients treated with Resveratrol-Copper identifies differential genes and pathways. **A**. MDS plot showing clusters between R-Cu treated (Teal) and untreated samples (Red). **B**. Volcano plot of statistically significant differentially expressed genes. Red dots represent up-regulated genes with log2 (fold change) > 2 and p-adjusted < 0.05, and blue dots represent down-regulated genes with log2 (fold change) < −2 and p-adjusted < 0.05. **C – D.** Gene set enrichment analysis (GSEA) plots showing de-enrichment of metastasis pathways (**C**) and cellular metaprogramming and heterogeneity pathways (**D**) respectively in R-Cu samples compared to untreated samples. For each gene set, Normalised Enrichment Score (NES), nominal p-value, and False Discovery Rate (FDR) are shown on the plot.

#### Identification of differentially expressed genes and pathways

A total of 955 differential expressed genes (DEGs) were identified through DESeq2 analysis, of which 85 genes showed upregulation with a Log2FC>2 and padj<0.05 and 870 genes showed downregulation with a Log2FC<-2 and padj<0.05 (Fig. 2B and Table S4). Gene set enrichment analysis (GSEA) was conducted to identify significantly enriched pathways in R-Cu treated samples. Notably, the upregulated genes in R-Cu treated samples included pathway involved in the insulin signaling (IGF2, IGFBP7), which is known to have tumor suppressor role in cancer by inducing apoptosis and senescence in solid tumors (Singh et al., 2020) (Fig. 2B). We also observed upregulation of extracellular matrix and related pathways (SERPINA5, CPA3, CCN3, IGFBP7, LEFTY2, SOD3 and F13A1) in R-Cu treated samples which play a role in role in modulating the extracellular matrix (Fig. 2B and Table S4). Upregulation of extracellular matrix pathways have been shown to prevent metastasis and improve patient survival (Bijsmans et al., 2011; Jing et al., 2014). The down-regulated pathways included intracellular ion channel genes (CFTR, CNGB1 and CNGB3) which modulate several of immunological and tumorigenic cellular processes (Peruzzo et al., 2016) (Fig. 2B and Table S4). Significant downregulation of pathways such as metastasis and CD8 T cell chromatin were also observed in R-Cu samples with NES<-1.5 and an FDR q-value<0.05 (Fig. 2C and 2D). Also significantly down-regulated with a log2FC of −22.77 and padj<0.05 was PVRIG-2P, a pseudogene of PVRIG, which is an evolutionary homologue of PD-L1 (Fig. 2B and Table S4) (Whelan et al., 2019; Yang et al., 2024).

#### Complex relationship between RNA and protein turn over

We found that cancer hallmark genes *BCL2, ATM*, and immune check point genes *CTLA4, KLRC1* (coding for NKG2A) showed downregulation in whole transcriptome analysis, which was also reflected in downregulation of their corresponding proteins (Figs. 1B, S7 and S2). However, we also observed discordant regulation of several hallmark, stem cell and immune check-point genes in our transcriptome analysis (Fig. S7 and Table S4). The hallmark genes *AKT1, IL6, MYC, NFKB1, TGFB1, TP53* (coding for pp53), *VEGFA, SLC2A1* (coding for GLUT1), *CDH2* (coding for N-cadherin), *CDKN21A* (coding for p21), *VIM* (coding for Vimentin); stem cell marker genes *CD44, SOX2* and immune check point genes *HAVCR2* (coding for TIM3), *LAG3* showed increased gene expression, while hallmark genes *RAD50, CCND1*, stem cell marker genes *PROM1* (CD133), immune check point genes *PDCD1* (coding for PD-1), *CD274* (coding for PD-L1) exhibited relatively unchanged expression levels (Fig. S7). On the other hand, at a protein level, their corresponding proteins uniformly exhibited downregulation (Figs. S2 & S3). Overall, an inverse correlation of gene expression and protein expression in 19 out of 24 analyzed cancer hallmarks, immune checkpoint, and stem cell markers indicates a complex regulatory relationship involving protein synthesis, protein degradation and / or post-translational modification.

#### R-Cu treatment increases intrinsic apoptotic cell death

A significant upregulation in gene set of hallmark apoptosis (NES = 2.899 and nominal p-value = 0) was observed in R-Cu treated samples suggesting intense activation of intrinsic cell death (Fig. 3A). Furthermore, proteasome degradation pathway, ubiquitin proteasomal system pathway and malignant metaprogram 8 proteasomal degradation pathway were also significantly upregulated in R-Cu treated samples (Figs. 3B, 3C and 3D). Fig. 3E provides a comprehensive perspective of the interconnecting network of genes involved in apoptosis and proteasomal degradation pathways. The data suggests that the combined activation of apoptosis and proteasomal degradation facilitates marked intrinsic cell death upon R-Cu treatment. Consistent with this interpretation is our IF results which showed Annexin V expression to be significantly reduced, suggesting that apoptotic cellular debris were being efficiently removed from GBM tumors via these pathways resulting in proteasomal degradation (Fig. S8). These data indicated that R-Cu treatment had led to an intense intrinsic apoptosis of GBM cells and that the apoptotic cellular debris was rapidly eliminated via proteosomal degradation resulting in our detection of a reduction in Annexin V in the R-Cu treated samples (Fig. S8).

**Figure 3:**
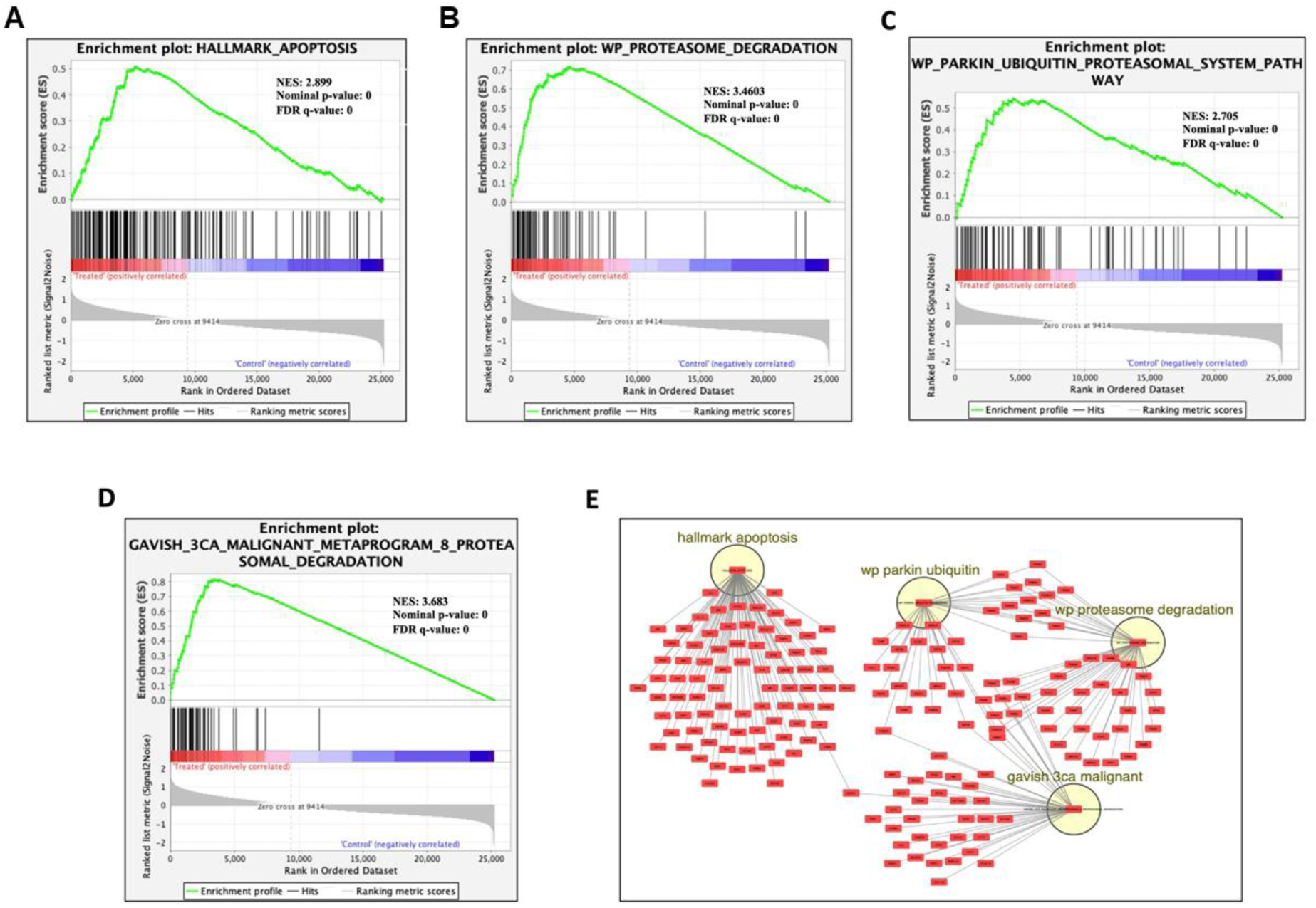
Transcriptomic signature indicates activation of intrinsic apoptosis and proteasomal degradation cascade upon R-Cu treatment. **A**. Enrichment plot generated using GSEA showing upregulation of hallmark apoptosis gene set, **B.** upregulation of proteosome degradation pathways, **C.** upregulation of Proteasomal System pathway, **D.** upregulation of Malignant Metaprogram 8 Proteasomal Degradation pathway, respectively in R-Cu treated samples. For each gene set, Normalised Enrichment Score (NES), nominal p-value, and False Discovery Rate (FDR) are shown on the plot. **E**. Network map showing genes involved in apoptosis and proteasomal degradation pathways.

## Discussion

The results of this study suggests that a simple, non-toxic and inexpensive combination of the commonly used nutraceuticals Resveratrol and Copper administered orally can have a profound effect on the aggressive phenotype of GBM. This effect was ostensibly mediated via oxygen radicals that were generated upon oral ingestion of R-Cu leading to deactivation of cfChPs in TME of GBM (Khanvilkar and Mittra, 2025; Mittra, 2024). Using whole transcriptome sequencing we detected activation of extensive apoptosis in the R-Cu treated samples. A marked reduction in Bcl-2 expression in the treated samples provided further evidence of activation of apoptosis of GBM cells following R-Cu treatment.

Immune staining followed by confocal microscopy of FFPE sections detected copious presence of cfChPs which had been released from the dying GBM cells in TME of control samples and which were virtually eliminated via the medium of ROS following a short course of R-Cu. These data indicated that R-Cu treatment had led to an intense intrinsic apoptosis of GBM cells and that the apoptotic debris was rapidly eliminated via proteosomal degradation. The latter correlated with our observation of an inverse relationship between protein expression and level of RNA transcription suggestive of efficient activation of proteasomal degradation in coordination with apoptosis (Abbas and Larisch, 2021; Yuan et al., 2022). We also observed a highly significant reduction in Ki-67, an indicator of higher tumour grade, to suggest that the tumours had been down-staged to a lower histological grade. Furthermore, significant down-regulation in hallmarks of cancer, immune check-points and stem cell markers suggested that aggressive behavior of GBM were markedly attenuated following a short course of R-Cu.

Whole transcriptome sequencing of R-Cu treated GBM samples detected upregulation of *IGFBP7* gene – a tumor suppressor known to inhibit proliferation and induce apoptosis and senescence (Singh et al., 2020). Furthermore, our detection of down-regulation of metastasis related genes such as *GATA2, PKD1, NFIA* suggestive of attenuation of aggressive behavior of GBM which aligns with independent reports in other cancer types (Chiang et al., 2014; Lee et al., 2014; Wang et al., 2015).

Immunotherapy of cancer is a major breakthrough in cancer treatment. However, these targeted therapies that are currently in use are expensive and often very toxic and are generally used one at a time. Against this backdrop, our results that a simple, inexpensive and non-toxic combination of R and Cu can simultaneously down-regulate six immune check-points is a highly significant finding. In this context, we have recently reported that cfChPs released from dying cancer cells that circulate in human blood, or those that are released from dying cancer cells, can activate immune checkpoints in human lymphocytes both *in vitro* and *in vivo* (Shabrish et al., 2024). The immune check-points *PD-1, CTLA-4, LAG-3, NKG2A*, and *TIM-3* were significantly activated when human lymphocytes were treated with cfChPs that were derived from cancer patient sera. The addition of R-Cu to the culture medium led to down-regulation of all five immune check-points. These results were confirmed in vivo in mouse splenocytes upon intravenous injection of cfChPs (Shabrish et al., 2024). These findings taken together with our present results would indicate that cfChPs are global instigators of immune check-points which can be down-regulated by treatment with the pro-oxidant combination of R and Cu. Thus R-Cu may offer a viable alternative to immunotherapy currently in practice in the treatment of cancer.

Our present results confirm those of our earlier exploratory study in advanced oral cancer which was primarily undertaken to determine the optimal dose for R and Cu (Pilankar et al., 2022). That study, conducted prior to the availability of R-Cu tablets, used oral administration of water-based suspensions of R and Cu and the optimum dose of R and Cu were adjudged to be 5.6 mg and 560 ng respectively. The bi-layered tablets used in the current study were developed on the basis of the result of this earlier study. Nonetheless, our earlier study showed that oral administration of the combination of R and Cu for 2 weeks could down-regulate multiple hallmarks of cancer and immune check-points in advanced oral cancer (Pilankar et al., 2022). The results of the earlier study and the current one suggest that cfChPs that are released from dying cancer cells into TME are global instigators for cancer hallmarks and immune checkpoints in the surviving cancer cells and are primarily responsible for the potentially lethal behavior of GBM.

In summary, results of our study suggest that oral administration of a non-toxic combination of small quantities of the commonly used nutraceuticals R and Cu can have a profound effect in attenuating the aggressive phenotype of GBM. They also suggest that cfChPs are the key instigators of cancer hallmarks, immune checkpoints and cancer stemness in GBM. Further studies are required to investigate whether prolonged treatment with R-Cu may induce the tumour to adopt a benign phenotype.

## Patients and Methods

### Ethics approval

This study was approved by Institutional Ethics Committee (IEC) (Approval no. 3456). All participants signed a written informed consent forms which were approved by the IEC.

### Patient details

Demographic information on the ten control and ten Resveratrol and Copper (R-Cu) treated patients is given in Table S1. All tumour tissues were histologically verified by a senior neuropathologist to be GBM (as per WHO 2021 classification).

### R-Cu tablets and dosage schedule

Bi-layer tablets containing 5.6 mg of R and 560 ng of Cu were provided by Inventia Healthcare Limited which were administered to GBM patients four times a day on an empty stomach. Control patient did not receive R-Cu tablets.

### Sample collection

Biopsy samples of the tumour were collected at operation. One half of the sample was placed in formalin for immunofluorescence (IF) analysis and the other half was placed in RNAlater for transcriptome sequencing.

### Fluorescence immune-staining and confocal microscopy

Fluorescence immune-staining using anti-DNA and anti-histone H4 antibodies followed by confocal microscopy was performed on formalin fixed paraffin embedded (FFPE) sections to detect the presence of cfChPs in TME of tumour tissues. The detailed protocol of the procedure is described by us earlier (Mittra et al., 2020b). Fluorescence intensity of five randomly chosen confocal fields (∼ 50 cells in each field) was recorded, and mean fluorescence intensity (MFI) (± S.E.M) was estimated.

### Immunofluorescence

#### Methodology

Indirect IF analysis on FFPE sections of tumour tissues was performed using a protocol described by us earlier (Pilankar et al., 2022). The various biomarkers analyzed by IF are listed in Table S2. The table also provides information on the sources and catalogue numbers of the primary and the secondary antibodies used for IF analysis. One thousand cells were analysed on each slide and the percentage of cells positive for fluorescent signals of the various biomarkers was recorded. The only exception was Annexin V where MFI per cell was recorded after analyzing one thousand cells.

#### Blinded analysis

IF analysis was done in a blinded fashion such that the examiner was unaware of the identity of the slides being analysed.

### Transcriptome analysis

#### Differential expression analysis

Raw RNA-seq FASTQ files were evaluated for quality control checks using FastQC tool (v0.12.1) (Andrews, 2010). Low-quality reads and adapter sequences were trimmed using Fastp (Chen et al., 2018) and the results of FastQC reports were summarized using multiQC (v1.15) (Ewels et al., 2016). Trimmed reads were then aligned to the Human GRCh38 transcriptome and Ensembl annotation v104 using Salmon (v1.5.1) (Patro et al., 2017) in a quasi-mapping-based mode, which enabled accurate transcript abundance quantification with GC bias and sequence level bias corrections. The transcript-level abundances were converted into gene-level counts using tximport (v1.18.0) package in R (Soneson et al., 2016). Differential gene expression analysis was performed using DESeq2 (v1.30.1) (Love et al., 2014) with Wald’s test for significance testing. The threshold for differential expression is set at log2fold-change (log2FC) of ±2 and adjusted p-value (padj)<0.05. These differential expressed genes (DEGs) were then visualised with a volcano plot created using EnhancedVolcano (Blighe, Kevin, Sharmila Rana, 2019).

#### Pathway Enrichment Analysis

Pathway enrichment analysis was performed using Gene set enrichment analysis (GSEA; v4.3.3) (Subramanian et al., 2005) on DESeq2-normalized counts, utilising msigdb.v2023.2.Hs.symbols.gmt gene set. The analysis involved 1000 phenotype permutations and used Signal2Noise ranking metric to generate the ranked list of genes. Enrichment was evaluated based on normalised enrichment score (NES), with positive scores indicating upregulation and negative scores indicating down-regulation. A false discovery rate (FDR) q-value≤0.05 was considered statistically significant. Additionally, the genes associated with apoptosis and proteasomal degradation pathways were visualized as a network map in Cytoscape (Shannon et al., 2003). AutoAnnotate, a Cytoscape app was used to enhance visualization by labelling and grouping the pathways, thus identifying distinct functional clusters (Kucera et al., 2016).

### Statistical analysis

Statistical analysis was performed using two-tailed unpaired student’s *t*-tests using GraphPad Prism 8 (GraphPad Software RRID:SCR_002798, Boston, MA, USA) and the results are expressed as mean ± SEM. The significance thresholds were: **\*** p<0.05, **\*\*** p<0.01, *** p<0.005, and **** p<0.0001.

## Funding

This study was supported by the Department of Atomic Energy, Government of India, through grant CTCTMC to the Tata Memorial Centre awarded to IM.

## Ethics

This study was approved by Institutional Ethics Committee (IEC) (Approval no. 3456). All participants signed a written informed consent forms which were approved by the IEC.

## Conflict of Interest

*The authors declare no competing interests*.

## Authors’ Contributions

**C. Bandiwadekar:** Data curation, formal analysis, investigation, visualization, methodology. **N. L. Devi:** Data curation, formal analysis, investigation, visualization, methodology, writing-original draft. **A.V. Moiyadi:** Conceptualization, data curation. **V. Singh:** Conceptualization, data curation. **P. Shetty:** Conceptualization, data curation. **S. Epari:** Data curation, methodology, writing-original draft. **H. Tandel:** Data curation, methodology. **R. Yelukar:** Data curation, investigation, methodology. **D. Poojary:** Data curation, investigation, methodology. **G. V. Raghuram:** Data curation, formal analysis, supervision, validation, methodology, project administration. **S. Shabrish:** Resources, data curation, formal analysis, supervision, validation, visualization, methodology, writing-original draft, project administration. **P. Chandrani:** Conceptualization, resources, formal analysis, supervision, validation, writing-original draft, project administration, writing-review and editing. **I. Mittra:** Conceptualization, resources, formal analysis, supervision, funding acquisition, validation, visualization, writing-original draft, project administration, writing-review and editing

## Data Availability

All data shown in the manuscript is available in supplementary. Raw NGS data is being submitted to SRA. The Accession number will be updated once it is quality checked by SRA. Any additional data will be made available upon reasonable request. All relevant raw data will be freely available to any researcher for noncommercial purposes on request.

## Acknowledgement

We thank Ms. Ruchi Joshi for her help with some of the IF experiments and Mr. Ashish Pawar for his help in preparing the manuscript.

## Notes

### Competing Interest Statement

The authors have declared no competing interest.

### Clinical Trial

CTRI/2020/10/028476

### Author Declarations

This study was approved by Institutional Ethics Committee (IEC) of Advanced Centre for Treatment, Research and Education in Cancer (ACTREC), Tata Memorial Centre (TMC) (Approval no. 3456). All participants signed a written informed consent forms which were approved by the IEC.

### Summary of Updates

We have made the following changes: 1) Changed the title of the article. 2) Added two new authors Ms. Roohi Yelukar and Ms. Disha Poojary. 3) Added a graphical abstract. 4) Changed the format of the references to comply with the requirements of eLife. 5) Made minor changes in the text.

