## Supplementary Information for "Downregulation of cancer hallmarks and immune check-points in patients with glioblastoma following a short course of the pro-oxidant combination of Resveratrol and Copper"

#### Figure S1

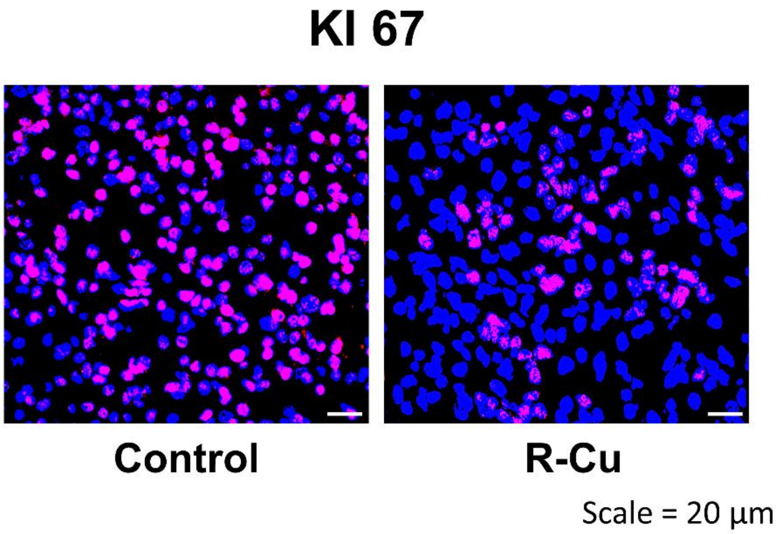


**Figure S1: Representative IF images of Ki-67 in control and R-Cu treated samples.** Quantitative histograms are presented in figure 1B.

**Figure S2**

#### Cancer hallmarks

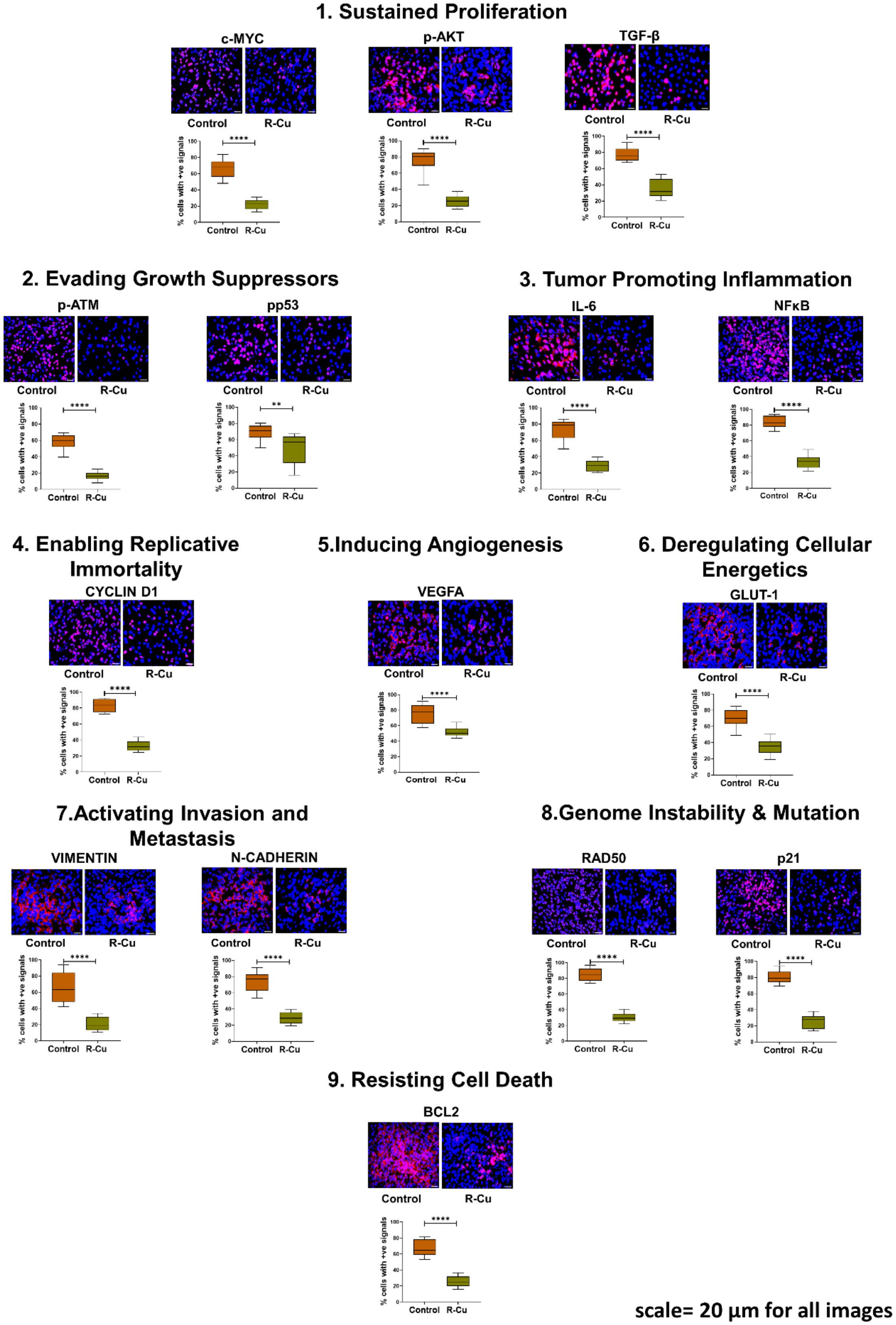


scale= 20 µm for all images

**Figure S2: Representative IF images of 16 biomarkers representing 9 hallmarks of cancer and quantitative results represented as boxplots.** n = 10 each in control and R-Cu groups. ** = $p < 0.01$; *** = $p < 0.001$; **** = $p < 0.0001$.

#### Figure S3

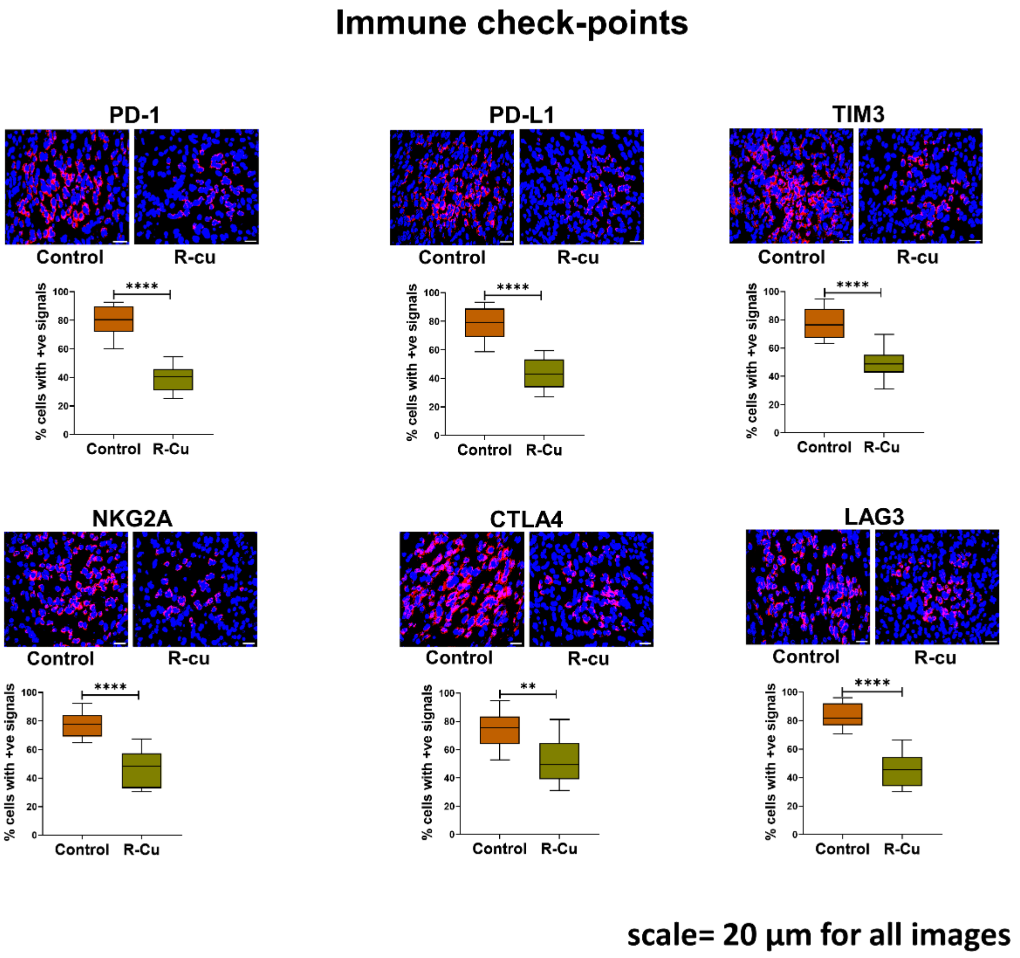


**Figure S3: Representative IF images of 6 immune check-points and quantitative results represented as boxplots.** n = 10 each in control and R-Cu groups. ** = $p < 0.01$; **** = $p < 0.0001$.

#### Figure S4

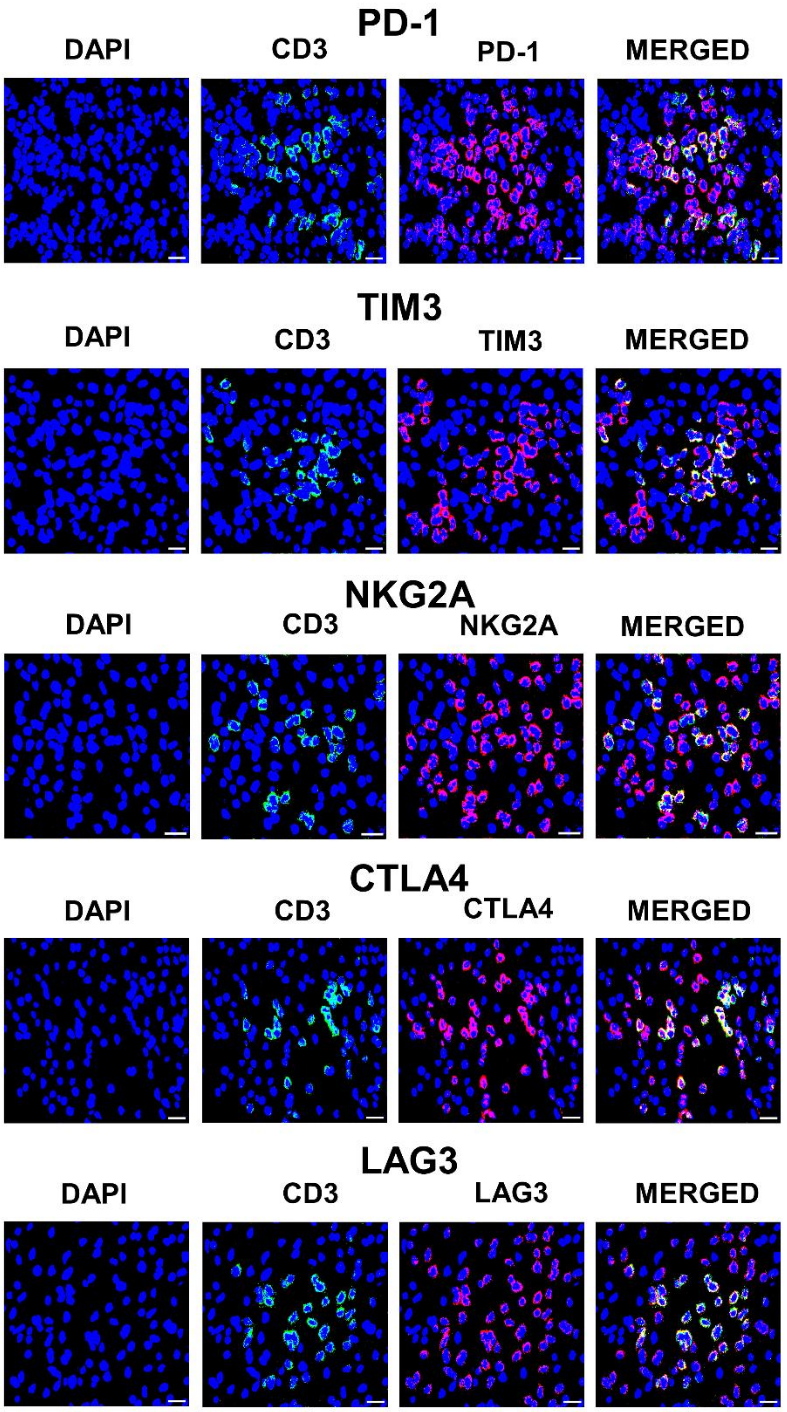


scale= 20 µm for all images

**Figure S4: Representative IF images showing co-expression of various immune checkpoints by tumour infiltrating lymphocytes represented by CD3.** FFPE sections of GBM tumour tissues were simultaneously immune-stained with antibodies against various immune-checkpoints and that against CD3 lymphocytes. Co-localisation of all five immune check-points and CD3 lymphocytes is clearly seen.

### Figure S5

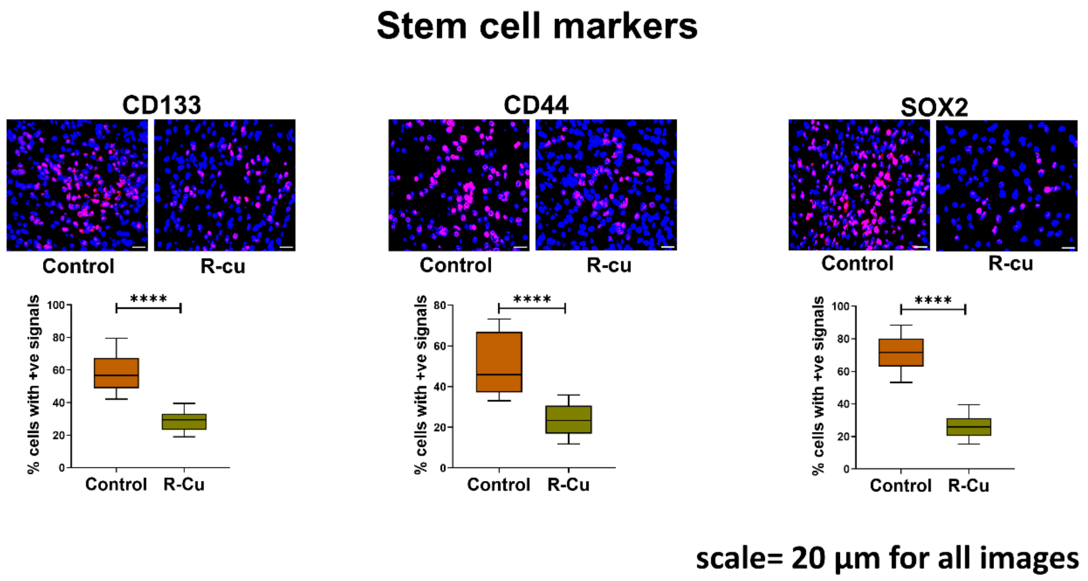


**Figure S5: Representative IF images of 3 stem cell markers and quantitative results represented as boxplots.** n = 10 each in control and R-Cu groups. **** = $p < 0.0001$.

#### Figure S6

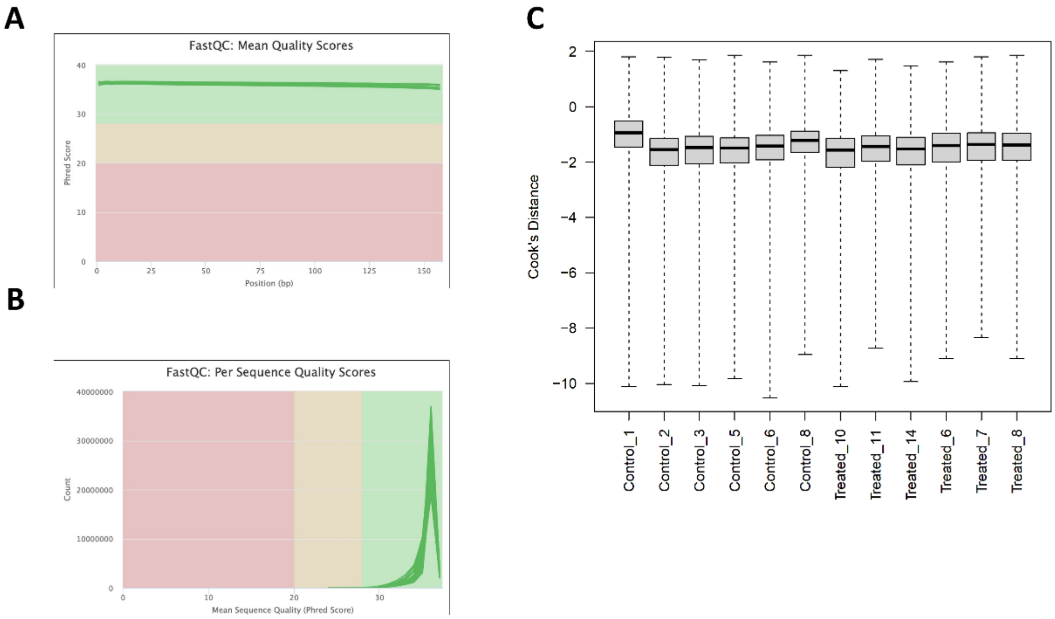


**Figure S6: RNA-seq quality check and normalization. A & B** FastQC plots showing mean per-base quality scores of RNA-seq data **C.** Cook's distance boxplot to visualize the data distribution post normalization

### Figure S7

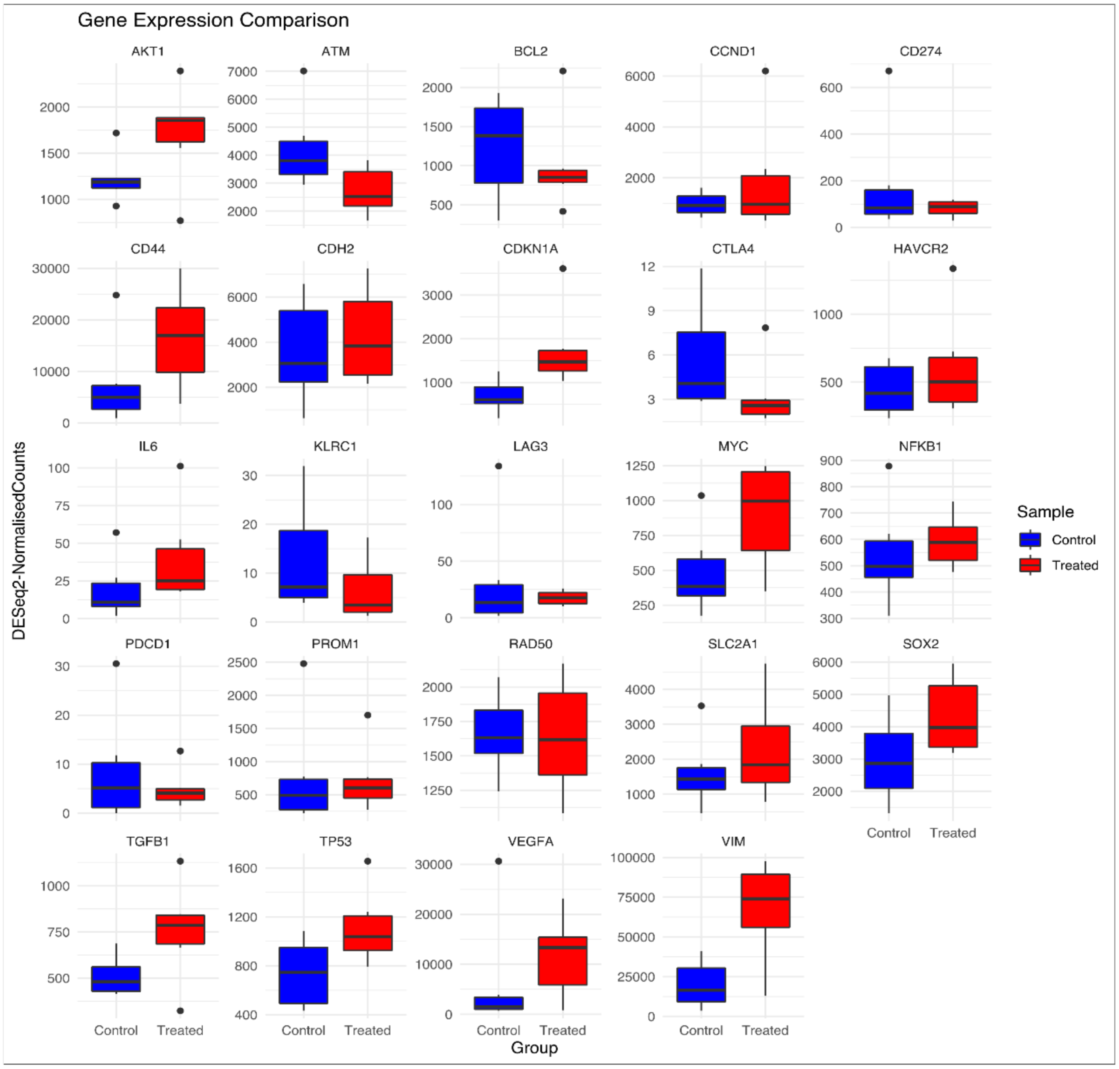


**Figure S7**: **Gene-wise comparison of expression in R-Cu treated and untreated samples.** Box plot representation of the selected cancer related genes

### Figure S8

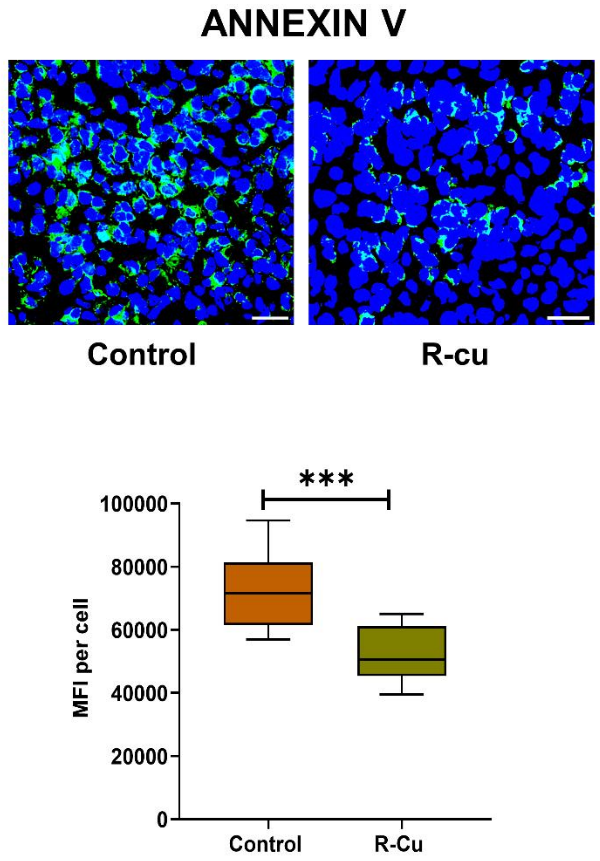


**Figure S8: Representative IF images of Annexin V in control and R-Cu treated samples.** Quantitative results are presented as boxplots. n = 10 each in control and R-Cu treated groups. *** = $p < 0.001$.

**Table S1:** Demographic information of control and Resveratrol and Copper (R-Cu) treated patients.

**Table S2:** List of biomarkers and antibodies used in this study.

**Table S3 & S4:** General information on RNA-seq data and a detailed list of differentially expressed genes
