## Supplementary material for "Downregulation of cancer hallmarks and immune check-points in patients with glioblastoma following a short course of the pro-oxidant combination of Resveratrol and Copper": Table S1

**Patient details**

**Control patients	R-CU patients**

| **SR.NO** | **AGE** | **SEX** |
| --- | --- | --- |
| 1 | 36-40 | M |
| 2 | 71-75 | M |
| 3 | 41-45 | M |
| 4 | 61-65 | M |
| 5 | 71-75 | M |
| 6 | 56-60 | M |
| 7 | 61-65 | M |
| 8 | 61-65 | F |
| 9 | 36-40 | M |
| 10 | 66-70 | M |

| **SR.NO** | **AGE** | **SEX** | **Pre op Res Cu (no. of days)** |
| --- | --- | --- | --- |
| 1 | 41-45 | F | 7 |
| 2 | 56-60 | M | 9 |
| 3 | 51-55 | M | 9 |
| 4 | 41-45 | F | 24 |
| 5 | 61-65 | M | 7 |
| 6 | 61-65 | M | 8 |
| 7 | 61-65 | M | 10 |
| 8 | 71-75 | M | 11 |
| 9 | 51-55 | M | 17 |
| 10 | 71-75 | M | 14 |

**Pre op Res Cu no. of days**

**11.6 ± 5.37 (mean ± SD)**
