## Supplementary material for "Downregulation of cancer hallmarks and immune check-points in patients with glioblastoma following a short course of the pro-oxidant combination of Resveratrol and Copper": Table S2

Cancer grading marker

|  | **Bio-marker** |
| --- | --- |
| **cancer grading marker** | Ki-67 |

Cancer hallmarks and corresponding biomarkers

| **Hallmark** | **Bio-marker** |
| --- | --- |
| **Sustained Proliferation** | TGF-β  c-Myc  pAKT |
| **Evading growth suppressor** | pATM  pp53 |
| **Tumor promoting inflammation** | IL-6  NF-κβ |
| **Enabling replicative immortality** | Cyclin D1 |
| **Inducing angiogenesis** | VEGFA |
| **Deregulating cellular energetics** | GLUT1 |
| **Resisting Cell Death** | Bcl-2 |
| **Genome instability and mutation** | Rad50  p21 |
| **Activating invasion and metastasis** | Vimentin  N-cadherin |
| **Avoiding immune destruction** | PD-1  PD-L1  CTLA-4  NKG2A  TIM-3  LAG3 |

Immune check-points and T-cell markers

|  | **Bio-marker** |
| --- | --- |
| **Avoiding immune destruction** | PD-1  PD-L1  TIM-3  NKG2A  CTLA-4  LAG3 |
| **T-cell marker** | CD3 |

Stem Cell markers

|  | **Bio-marker** |
| --- | --- |
| **Stem Cell markers** | CD133  CD44  SOX2 |

**List of antibodies used:**

Primary antibody for cancer grading

| **Biomarker** | **Catalogue No.** | **Company** |
| --- | --- | --- |
| Ki-67 | 556003 | BD Biosciences, USA |

Primary antibodies for cancer hallmark biomarkers:

| **Hallmark** | **Biomarker** | **Catalogue No.** | **Company** |
| --- | --- | --- | --- |
| **Sustained Proliferation** | TGF-β | A92358 | Antibodies.com, UK |
|  | c-Myc | 5605S | Cell Signalling Technologies, USA |
|  | p-Akt | 4060S | Cell Signalling Technologies, USA |
| **Evading Growth Suppressors** | pp53 | 700439 | Invitrogen -Thermo-Fisher Scientific, USA |
|  | p-ATM | 05-740 | Merck-Sigma-Aldrich, Germany |
| **Avoiding Immune Destruction** | PDL-1 | A93289 | Antibodies.com, UK |
|  | PD-1 | 86163S | Cell Signalling Technologies, USA |
|  | CTLA-4 | A54651 | Antibodies.com, UK |
|  | NKG2A | A39318 | Antibodies.com, UK |
|  | TIM-3 | A84174 | Antibodies.com, UK |
|  | LAG-3 | ab209236 | Abcam, UK |
| **Enabling Replicative Immortality** | CCND-1 | sc-246 | Santa-Cruz Biotechnology, USA |
| **Tumor Promoting Inflammation** | IL-6 | ab6672 | Abcam, UK |
|  | NF-ĸβ | 8242S | Cell Signalling Technologies, USA |
| **Activating invasion and metastasis** | Vimentin | 5741S | Cell Signalling Technologies, USA |
|  | N-cadherin | ab76057 | Abcam, UK |
| **Inducing Angiogenesis** | VEGFA | ab 52917 | Abcam, UK |
| **Genome Instability and Mutation** | p21 | sc-817 | Santa Cruz Biotechnology, USA |
|  | Rad50 | sc-20155 | Santa Cruz Biotechnology, USA |
| **Resisting Cell Death** | Bcl-2 | ab692 | Abcam, UK |
| **Deregulating Cellular Energetics** | Glut-1 | ab652 | Abcam, UK |

Primary antibodies for immune check-points and T-cell marker

|  | **Biomarker** | **Catalogue No.** | **Company / vendor** |
| --- | --- | --- | --- |
| **Avoiding Immune Destruction** | PDL-1 | A93289 | Antibodies.com, UK |
|  | PD-1 | 86163S | Cell Signaling Technologies, USA |
|  | CTLA-4 | A54651 | Antibodies.com, UK |
|  | NKG2A | A39318 | Antibodies.com, UK |
|  | Tim-3 | A84174 | Antibodies.com, UK |
|  | LAG-3 | ab209236 | Abcam, UK |
| **T-cell marker** | Anti-Human CD3-FITC | 561807 | BD Biosciences, USA |

Primary antibodies for stem cell markers:

| **Sr. No** | **Antibody** | **Catalogue No.** | **Company/Vendor** |
| --- | --- | --- | --- |
| 1. | CD133 | 64326S | Cell Signaling Technologies, USA |
| 2. | CD44 | 3570S | Cell Signaling Technologies, USA |
| 3. | SOX2 | 14962S | Cell Signaling Technologies, USA |

Antibodies against DNA and Histone H4

| **Sr. No** | **Antibody** | **Catalogue No.** | **Company/Vendor** |
| --- | --- | --- | --- |
| 1. | Anti-DNA | NB110-89473 | Novus Biologicals LLC, USA |
| 2. | Anti-Histone H4 | Custom-synthesised | Bioklone Biotech Pvt Ltd., India |

Secondary antibodies used:

| **Sr. No** | **Antibody** | **Catalogue No.** | **Company/Vendor** |
| --- | --- | --- | --- |
| 1. | Donkey anti-Goat IgG (H&L) Texas Red | ab6883 | Abcam, UK |
| 2. | Goat anti-Mouse IgG (H&L) TRITC | ab6786 | Abcam, UK |
| 3. | Goat anti- Rabbit IgG (H&L) TRITC | ab6718 | Abcam, UK |
| 4. | Goat anti-Rabbit IgG FITC | AP307F | Merck-Sigma-Aldrich, Germany |
